# The toll of COVID-19 on African children: A descriptive analysis on the COVID-19-related morbidity and mortality among the pediatric population in Sub-Saharan Africa

**DOI:** 10.1101/2021.07.02.21259857

**Authors:** Sabina Rodriguez Velásquez, Léa Jacques, Jyoti Dalal, Paolo Sestito, Zahra Habibi, Akarsh Venkatasubramanian, Benedict Nguimbis, Sara Botero Mesa, Cleophas Chimbetete, Olivia Keiser, Benido Impouma, Franck Mboussou, George Sie William, Nsenga Ngoy, Ambrose Talisuna, Abdou Salam Gueye, Cristina Barroso Hofer, Joseph Waogodo Cabore

**Author notes:** These authors contributed equally to the work (Cristina Barroso Hofer, Joseph Waogodo Cabore).

## Abstract

**Introduction:** Since the beginning of the COVID-19 pandemic, very little data on the epidemiological characteristics among the pediatric population in Africa has been published. This paper examines the age and sex distribution of the morbidity and mortality rate in children with COVID-19 and compares it to the adult population within 15 Sub-Saharan African countries.

**Methods:** A merge line listing dataset using a reverse engineering model shared by countries within the Regional Office for Africa was analyzed. Patients diagnosed within 1 March 2020 and 1 September 2020 with confirmed positive RT-PCR test for SARS-CoV-2 were analyzed. Children’s data were stratified into three age groups: 0-4 years, 5-11 years, and 12-17 years, while adults were combined. The cumulative incidence of cases including its medians and 95% confidence intervals were calculated.

**Results:** 9% of the total confirmed cases and 2.4% of the reported deaths were pediatric cases. The 12-17 age group in all 15 countries showed the highest cumulative incidence proportion in children. COVID-19 cases in males and females under the age of 18 were evenly distributed. Among adults, a higher case incidence per 100,000 people was observed compared to children.

**Conclusion:** The cases and deaths within the children’s population was smaller than the adult population. These differences can reflect biases in COVID-19 testing protocols and reporting implemented by countries, highlighting the need for more extensive investigation and focus on the effects of COVID-19 in children.

## Introduction

Since its emergence in December 2019 in the Chinese city of Wuhan, the coronavirus disease 2019 (COVID-19) caused by the severe acute respiratory syndrome coronavirus 2 (SARS-CoV-2) has infected millions of people around the world and caused over 3.9 million officially registered deaths (WHO, 2021). From the beginning of this pandemic, the pathogenic aspects in the pediatric population have remained less clear, especially in Sub-Saharan Africa where specialists’ initial projections estimated a high number of cases and deaths (Cabore et al., 2020). At the beginning of the pandemic, while schools were often closed, the role of children on SARS-CoV-2 transmission was often unclear as few disaggregated data on the infection and lethality rate existed. Nevertheless, as the pandemic progressed, more evidence regarding the transmission of SARS-CoV-2 in children became available and it became clear that children acquired COVID-19 possibly at a similar rate than adults while being a possible source of further transmission in households (Laws et al., 2021). Children were often reported to have mild, non-specific symptoms such as fever and mild respiratory symptoms and rarely present severe symptoms such as pneumonia (Idele et al., 2020, Ludvigsson, 2020, Yoon et al., 2020). Some of the severe cases have been attributed to a new medical entity named multisystem inflammatory syndrome in children (MIS-C), which is defined as an acute inflammation affecting several organs and systems in the body. Although children with MIS-C often required intensive care, their mortality rates remained relatively low (Hoste et al., 2021). When comparing the proportion of deaths and mortality rate of children against adults, the adult population experienced higher numbers, globally. This phenomenon could be explained by differences in gene expression and comorbidities (Balasubramanian et al., 2020, Felsenstein and Hedrich, 2020, Hedrich, 2020). Moreover, the few age-specific SARS-CoV-2 infection fatality rate (IFR) studies that were conducted thus far tend to show an IFR close to 0% in young populations (Levin et al., 2020, Perez-Saez et al., 2021). In this way, African countries could benefit from an overall younger population to counterbalance the limits of their health system capacities (Ghisolfi et al., 2020, Walker et al., 2020, Zar et al., 2020).

As funds were massively redirected from supporting programs, fear for children’s disease outcomes in African countries has grown; since the population under the age of 18 constitutes a higher part of society in low and middle-income countries (LMICs) and constitutes a considerable proportion of the population with comorbidities such as malnutrition, HIV, and other illnesses (Buonsenso et al., 2020, El-Sadr and Justman, 2020, Roberton et al., 2020, Zar et al., 2020). As Sub-Saharan countries show quite heterogeneous socio-economic contexts, high variation in the time-course and burden of COVID outbreaks can be expected (Cabore et al., 2020, Diop et al., 2020, Rice et al., 2021, Van Damme et al., 2020). Since very little data has been published about the epidemiological characteristics of COVID-19 among the pediatric population in Africa, the objective of this paper is to describe the morbidity and mortality in children in the African region. Specifically, the paper estimates the age and sex distribution of the morbidity and mortality rate in children with COVID-19 within African countries. Additionally, as one of the first studies of COVID-19 in African children, the paper aims to compare the number of diagnosed cases and mortality between children and adults with the objective of identifying the proportion of children cases compared to the total reported cases.

## Methods

### Study design

We conducted a retrospective study during the period 1 March and 1 September 2020. Our primary data source was the national situation reports made public by all countries experiencing the COVID-19 pandemic. The data from each situation reports were extracted and merged into a line listing of cases. Among the 47 member states of WHO AFRO, 34 provided publicly available data disaggregated by age and sex to WHO. The selection was further refined by excluding countries that did not have the data and variables of interest, resulting in a total of 15 countries meeting the inclusion criteria. Data from the following 15 countries were included in the analysis: Botswana, Burkina Faso, Chad, Congo, Eswatini, Guinea, Liberia, Mauritius, Mozambique, Namibia, Niger, Rwanda, Sao Tome e Principe, Sierra Leone, and Uganda. A more detailed flowchart and map for the selection of the countries in the analysis can be found in Figure 1 in the Supplementary Material. Since each country had different index case dates, we restricted the timeframe of the cases to include confirmed, real-time reverse transcription-polymerase chain reaction (RT-PCR) COVID-19 positive cases for SARS-CoV-2 diagnosed between 1 April 2020 and 1 September 2020. Laboratory confirmed cases were defined as a positive nucleic acid amplification test (NAAT) in the form of positive RT-PCR. All data were cleaned and analyzed centrally. Data from countries with only ≤35 percent missing values in age, sex, or clinical outcomes were included into our final sample population. Cases with missing values for laboratory results, age, and sex among the selected countries were removed from the sample while cases with missing values for clinical outcomes were kept in the analysis; nevertheless, only confirmed cases containing information on age, sex, and date of reporting were considered in the final sample size. Clinical outcomes were defined as alive or dead. Children were stratified into three age groups: 0-4 years (under-five), 5-11 (children), and 12-17 years (adolescents), while data from the adult population was combined.

**FIGURE 1.**
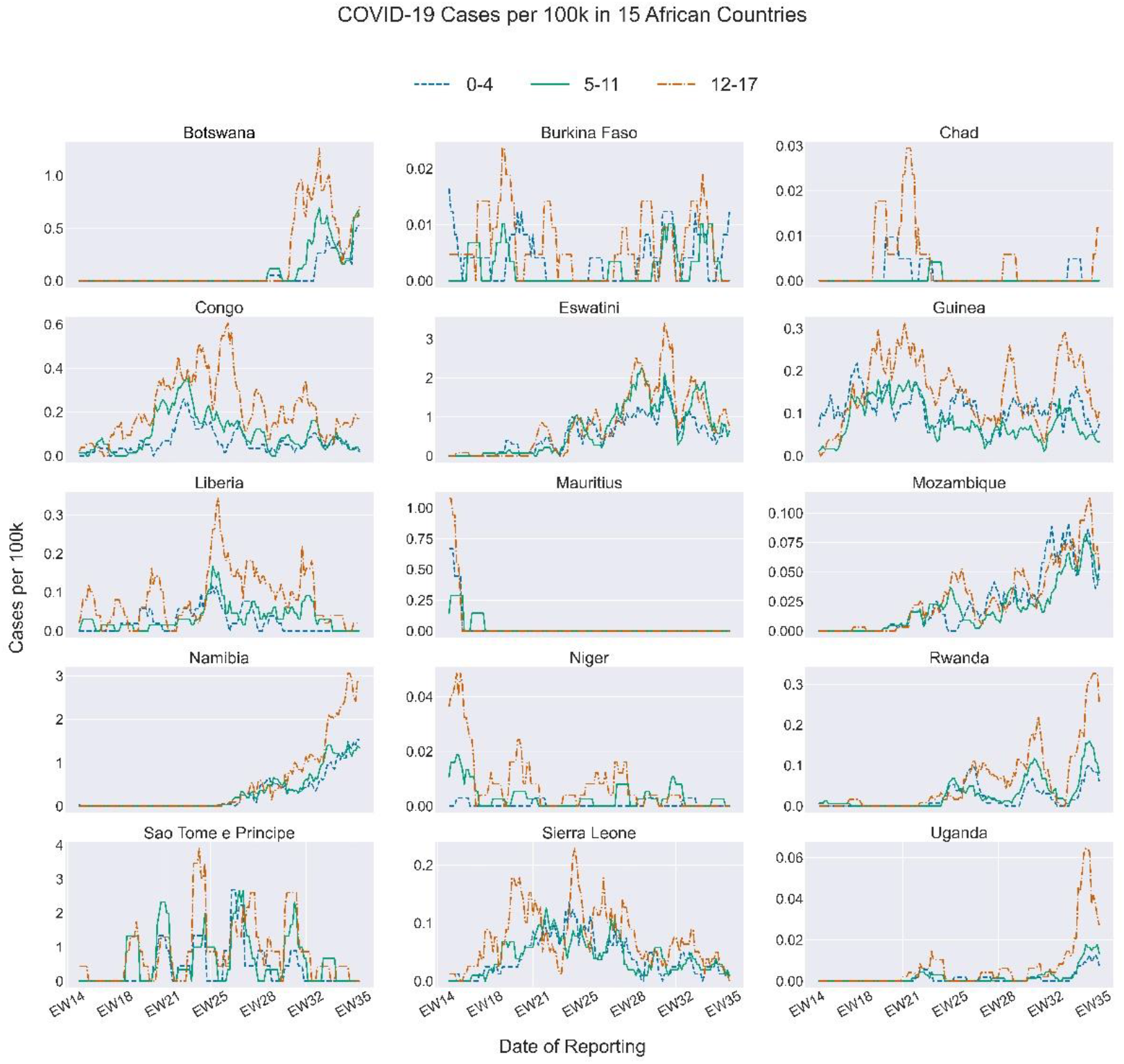
COVID-19 incidence among children aged 0-4 years, 5-11 years, and 12-17 years for 15 African countries, by epidemiological week - 1 April 2020 to 1 September 2020

### Outcomes and analysis

The case data for all COVID-19 positive cases were analyzed descriptively while focusing on the 0-to-17-year age groups. We calculated the cumulative incidence of cases, including its medians and 95% confidence intervals (CIs) by age, sex, and country. Wald’s method for binomial distribution was used when calculating the confidence interval. Similarly, the proportion of deaths and its median by age, sex, and country was calculated. For the calculation of incidence, the demographic information about total population by age, sex, and country was identified from the United Nations’ *World Population Prospects 2019*. Additionally, sub-regional groupings were created to facilitate the comparison of results within regions. This was performed by dividing the African continent into four principal sub-regional regions (West Africa, Central Africa, East Africa, and Southern Africa) and assigning each country into a sub-region according to its geographical location and the United Nations geoscheme for Africa. Trends in the number of cases were analyzed by calculating seven-day moving averages. Microsoft Excel, the R software (version 4.0.2) and Python (version 3.7.7) were used to perform the data analyses.

## Results

### General characteristics of COVID-19 cases

Table 1 details the characteristics of the cases included in the analysis, which contains 51,964 confirmed cases of SARS-CoV-2. Cases were more frequent in males (61.5%) than females. Pediatric cases accounted for 9.0% of reported cases, and the number of cases increased with age. Adolescents were the most affected age group in the pediatric population. Approximately 91% of the cases were reported in the adult population. A total of 1,110 deaths were reported from the positive SARS-CoV-2 cases (2.1% of the entire sample population); 27 deaths were reported in children <18 years of age (2.4% of dead cases). Based on the reported cases, Guinea, Congo, and Namibia were the countries with most positive SARS-CoV-2 cases accounting for 19.6%, 18.1%, and 14.4% of all the cases, respectively. African islands such as Mauritius and Sao Tomé e Principe reported 0.4% and 1.7% of the overall cases, respectively.

**TABLE 1.** Sample characteristics of population with COVID-19 (children and adults); 15 African countries, 1 September 2020.

| Variables | N = 51 964 | % |
| --- | --- | --- |
| **Sex** |  |  |
| Female | 19 996 | 38.5 |
| Male | 31 968 | 61.5 |
| **Age** |  |  |
| 0-4 | 1 146 | 2.2 |
| 5-11 | 1 505 | 2.9 |
| 12-17 | 2 025 | 3.9 |
| 17+ | 47 288 | 91.0 |
| **Clinical Outcome** |  |  |
| Alive | 50 192 | 96.5 |
| Dead | 1 110 | 2.1 |
| NA | 662 | 1.3 |
| **Country** |  |  |
| Botswana | 1 896 | 3.6 |
| Burkina Faso | 1 007 | 1.9 |
| Chad | 909 | 1.7 |
| Congo | 9 381 | 18.1 |
| Eswatini | 4 595 | 8.8 |
| Guinea | 10 198 | 19.6 |
| Liberia | 1 293 | 2.5 |
| Mauritius | 209 | 0.4 |
| Mozambique | 3 995 | 7.7 |
| Namibia | 7 485 | 14.4 |
| Niger | 1 081 | 2.1 |
| Rwanda | 4 035 | 7.8 |
| Sao Tome e Principe | 882 | 1.7 |
| Sierra Leone | 2 028 | 3.9 |
| Uganda | 2 970 | 5.7 |

As shown in Table 2, the total pediatric cumulative incidence per 100 000 people ranged from 0.2 in Chad to 103.5 in Eswatini. The median cumulative incidence per region increased with older age, for the exception of the Southern African sub-region which displayed the highest median cumulative incidence in the 5-11 age group. A total of 11 countries showed a positive association between age and total cumulative incidence, whereas 4 out of 15 countries showed a higher incidence among the age group 0-4 compared to age group 5-11 including Burkina Faso, Guinea, Chad, and Mozambique. Nevertheless. The 12-17 age group in all 15 countries showed the highest cumulative incidence proportion. Inter-age group variation ranged from 1:1.4 (Mozambique) to 1:16.7 (Chad). Cumulative incidence was highest in Central Africa with a median of 18.5 and one country listed in the top four highest cumulative incidences (Sao Tome e Principe). Southern Africa had the second highest median incidence of 16.9 with Western Africa (6.9) and Eastern Africa (5.6) following, respectively.

**TABLE 2.**
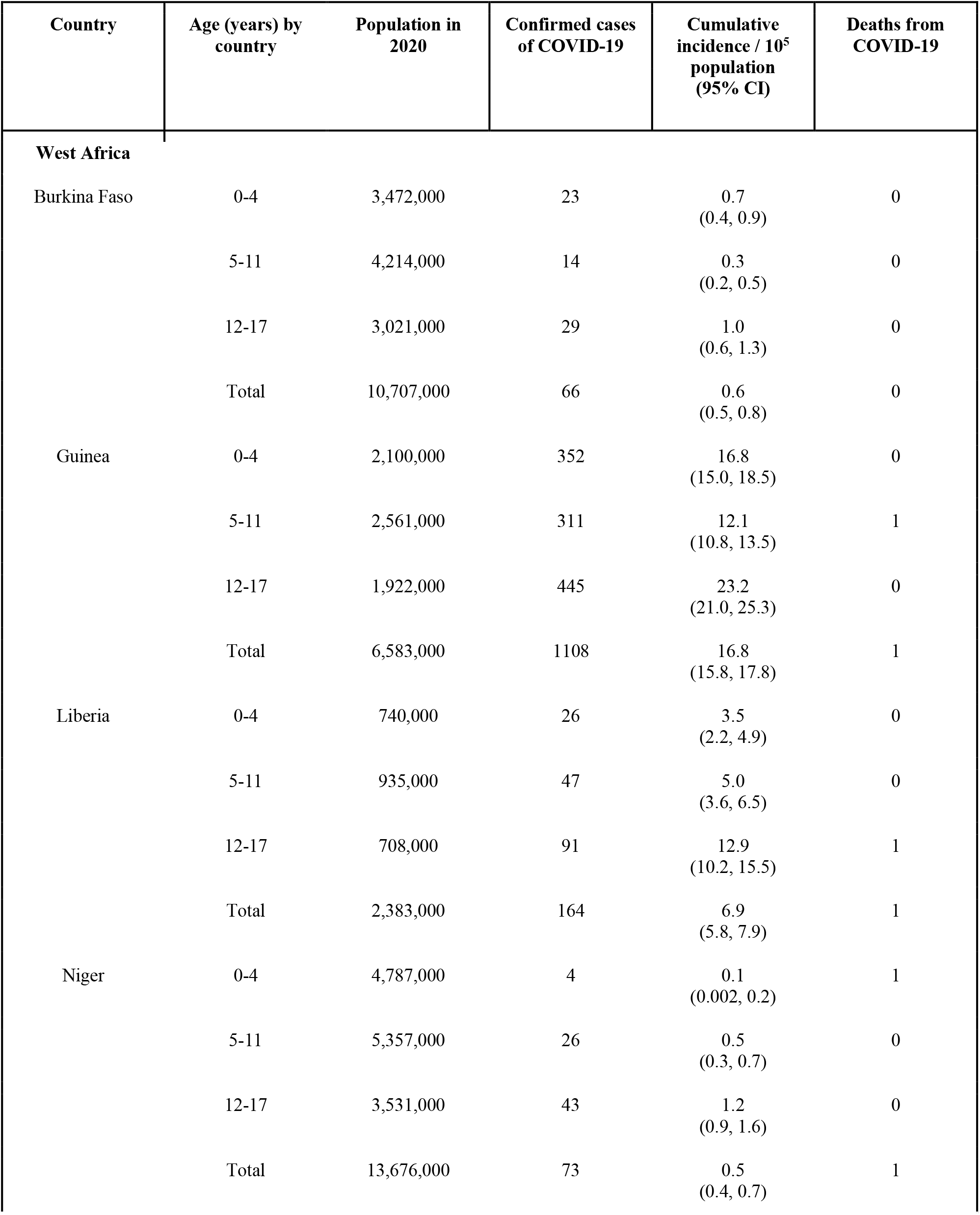
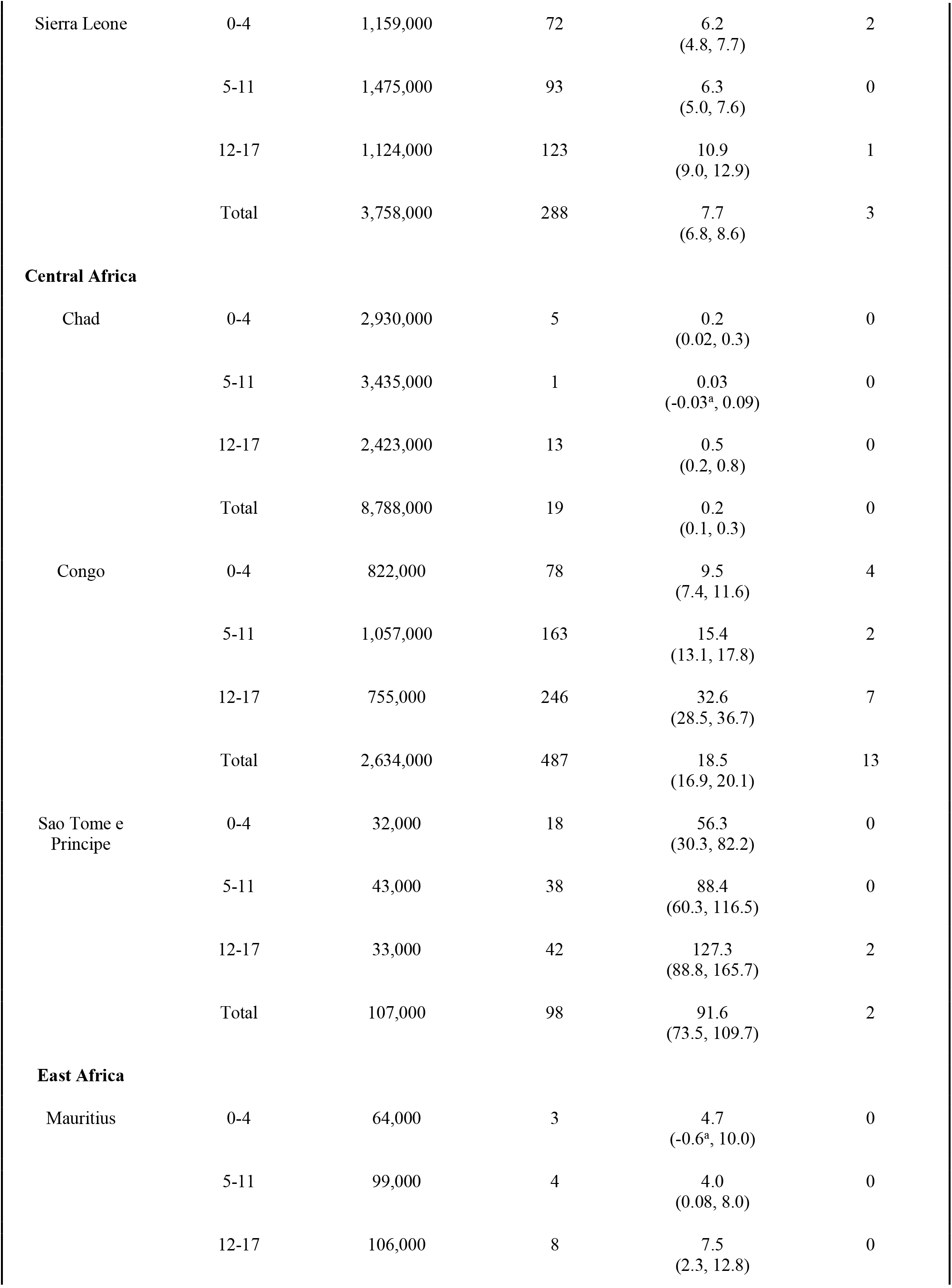
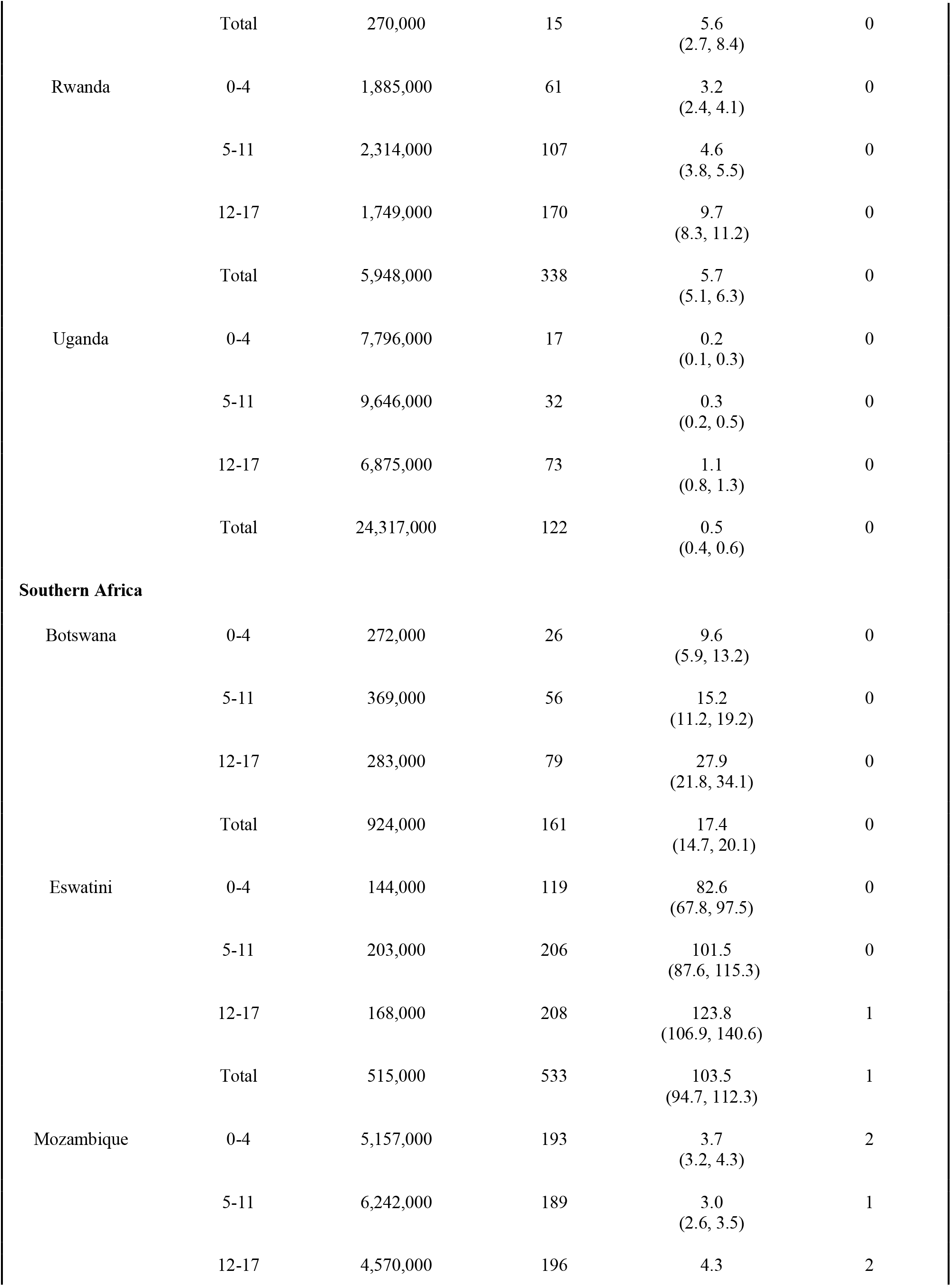
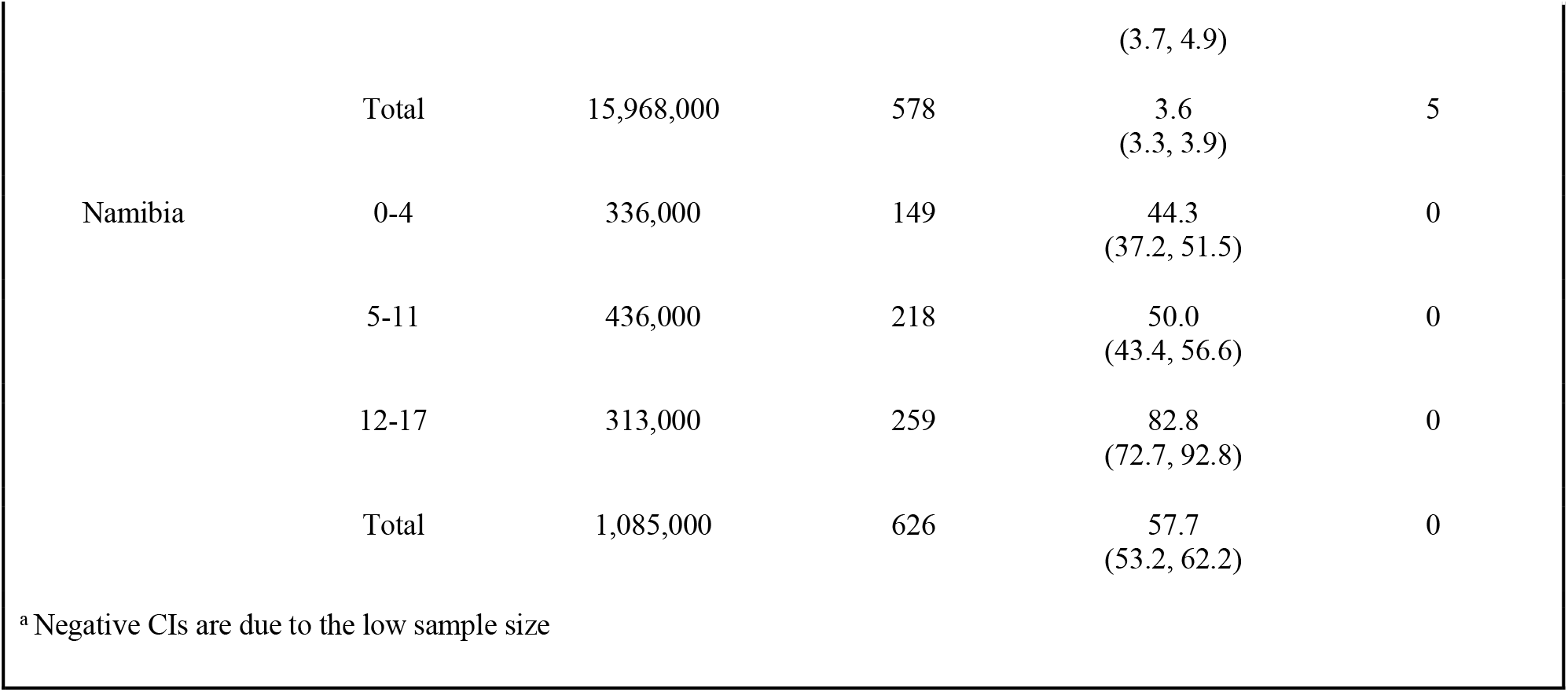
Distribution of pediatric cases, incidence, and deaths according to age groups; 15 African countries, 1 September 2020.

Mortality among children was relatively low as seven countries out of 15 did not report any deaths in children. Nevertheless, a total of 27 deaths were reported in eight different countries throughout Africa, of whom 15 were located in Central Africa, six in Western African, and six in Southern Africa. Over half of the recorded deaths (fourteen deaths) were recorded in adolescents aged 12 to 17 years. Children under five followed the adolescents in the number of deaths with nine overall reported deaths, while children aged five to eleven only reported a total of four deaths. Congo reported the most deaths (13) with the majority of fatal outcomes distributed among the 12 to 17 age group. Apart from Congo, Mozambique, Sierra Leone, and Sao Tomé e Principe reported five, three, and two deaths respectively, while Guinea, Liberia, Niger, and Eswatini only reported one death among those aged below 18 years.

Figure 1 shows the 15 epidemiological curves for all countries by age group. Ten countries including Chad, Congo, Eswatini, Guinea, Liberia, Mauritius, Mozambique, Niger, Sao Tomé e Principe, and Sierra Leone started reporting pediatric cases early on during the pandemic (Figure 1). In contrast, Botswana, Namibia, and Uganda did not report any COVID-19 cases in children until after the epidemiological week (EW) 21 (mid-May 2020). Although Rwanda reported cases early in April, the number of new cases reached 0 shortly after. Throughout June, case numbers, once again, increased in Rwanda, following a similar trend to Botswana, Namibia, and Uganda. Additionally, those four countries experienced their highest peak in cases around August 2020 with the majority of the peaks occurring between 19 July 2020 and 30 August 2020. Mauritius showed a unique trend in the number of pediatric cases over time. As noted in Figure 1, Mauritius experienced a high peak of cases at the beginning of April; however, after 26 April 2020, the number of cases plateaued until the end of August 2020. Some regional trends become evident while examining Figure 1. Southern African countries reported their first new children’s cases later in the pandemic than the West African countries. A similar trend can be examined for the Eastern African countries where the highest cases reported were found in epidemiological weeks 34 to 36 (mid-August to end of August). Mauritius recorded all cases at the beginning of April and then flattened the curve and reported zero cases by the end of April.

Figure 2 provides the overall epidemiological curve of children (0 to 17 years old) compared to adults (18 to 64 years old) in all 15 countries. At the beginning of the study, the average mean number of reported cases in all children under 18 for the epidemiological week 15 (April 5 to April 11) was 0.009 cases per 100,000 people. During epidemiological week 19 (week of 9 May 2020), the number of reported cases approximately doubled, increasing to 0.018 mean reported cases per 100,000 people. Throughout May, the mean reported number of cases in children ranged from 0.018 to 0.03 cases per 100,00 people with the lowest number occurring during the week of 16 May 2020 (EW 20) and the highest number occurring during the week of 30 May 2020 (EW22). The mean reported number of cases in children for June ranged from 0.025 to 0.032 cases per 100,000 people and 0.027 to 0.043 cases per 100,000 people for July. The mean number of cases for the month of August ranged from 0.041 to 0.064 cases per 100,000 people with the overall highest number of reported cases during the week of 29 August 2020 (EW35). In adults, the month of April ranged from 0.11 to 0.17 cases per 100,000 people. The following months of May, June, and July had a similar increase in the number of reported cases with May ranging from 0.21 to 0.29 cases per 100,000 people, June ranging from 0.28 to 0.34 cases per 100,000 people, and July ranging from 0.32 to 0.4 cases per 100,000 people. The number of reported cases in adults for the month of August ranged from 0.36 to 0.74 cases per 100,000 people.

**FIGURE 2.**
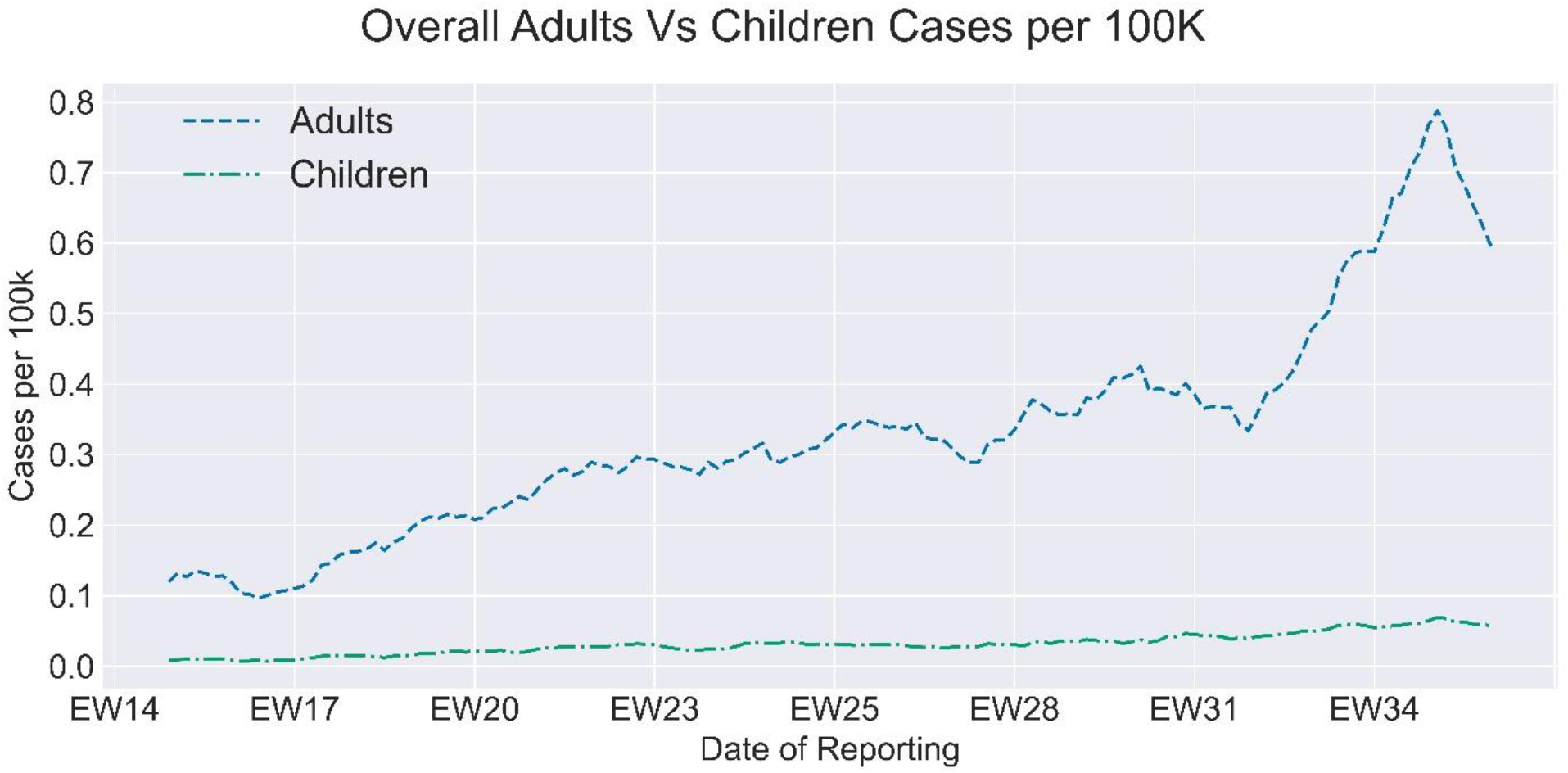
Overall COVID-19 incidence in children (0-17 years) versus adults (18-64) by epidemiological week - 1 April to 1 September.

Figure 3 shows the age-sex distribution of COVID-19 cases in children. Across all countries, a relatively even distribution of COVID-19 cases between males and females under the age of 18 can be noted with more than half of the selected countries (9 countries) reporting more cases in males than females. Nonetheless, Botswana, Burkina Faso, Congo, Eswatini, Namibia, and Sierra Leone were different as they reported more female cases in their children population than male cases. Comprehensively, the age-group with the highest difference in cases by sex is the 12-17 years old age group.

**FIGURE 3.**
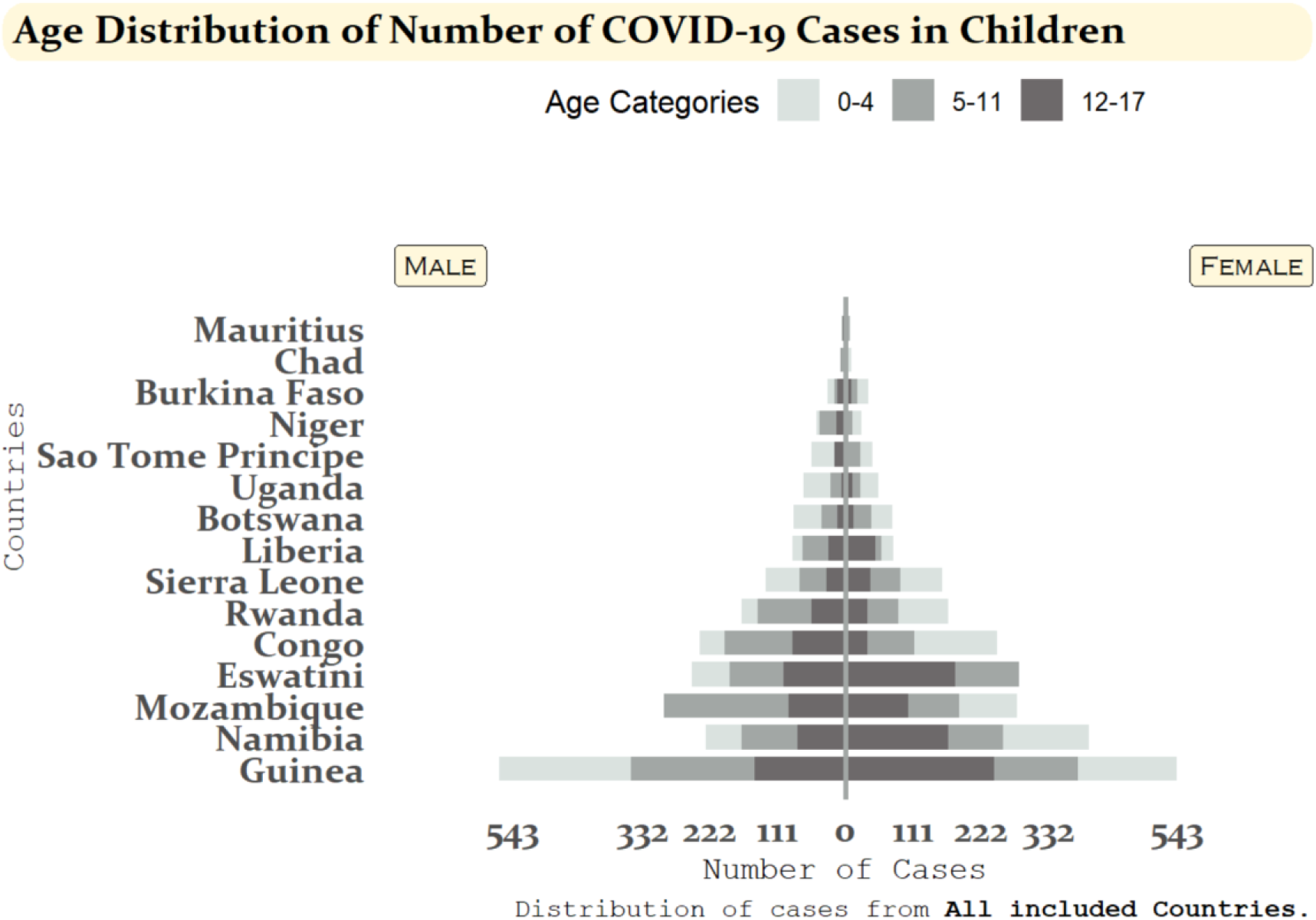
Age-sex distribution of COVID-19 cases and deaths among children aged 0-4 years, 5-11 years, and 12-17 years for 15 African countries: 1 April 2020 - 1 September 2020

## Discussion

### Major findings and trends

As one of the largest studies analyzing the number of cases and deaths caused by COVID-19 in Sub-Saharan Africa, our results show that the reported SARS-CoV-2 cumulative incidence proportions in the African pediatric population remained very low throughout the pandemic, and few deaths were attributed to COVID-19. Incidence proportions increased with age, whereas adolescents aged 12 to 17 years were the most affected pediatric group. Additionally, males tended to be slightly more affected than females throughout the selected countries. Globally, these results align with previous epidemiological studies. For instance, in a systematic review identifying 45 papers on COVID-19 in children, Ludvigsson et al. (2020) reported that children had accounted for 1-5% of COVID-19 cases and only one death throughout the entirety of articles analyzed (Ludvigsson, 2020). Additionally, no significant difference in the sex distribution of cases was observed. Interestingly, the largest Chinese pediatric case series cited in this review reported a median age of seven years, which differs from our findings. Among the few studies conducted in Africa providing individual-level data, a descriptive study from Nigeria showed trends that are comparable to our study (Elimian et al., 2020). In this study, children aged under 5, 5-13, and 14-20 years accounted for 1.7%, 3.9% and 6.1% of all confirmed cases respectively, a trend apparent in our study. Equally, males were more affected than females (65.8% vs 31.6%). The low number of diagnosed cases and the higher number of cases among older children may reflect age-dependent susceptibility to infection, milder or more asymptomatic cases occurring more frequently in younger children leading to less tests being performed, and differences in contact patterns between younger children and adolescents (Davies et al., 2020). Furthermore, under-diagnosis in children versus adolescents and adults may explain such differences.

As noted in Figure 2, the adult population experienced a higher number of cases per 100,000 people with a range of 0.110 to 0.75 reported COVID-19 cases per 100,000 people throughout time, whereas the children population reported relatively lower cases with their numbers not exceeding the 0.07 margin. Adults experienced three peaks in the number of cases with the first distinguishable peak occurring during the epidemiological week 25 (June 14), the second one occurring during the epidemiological week 31 (July 27), and the final major and highest one occurring around 29 August 2020. Similarly, children experienced their highest number of reported cases around 29 August 2020 (EW35). Nevertheless, adults had a constant increase in the number of reported cases throughout time, while the number of reported cases in children remained relatively similar throughout time. Other studies such as the one conducted in the United States comparing COVID-19 trends among different age groups, also reported higher incidence among adults than children (Leidman E, 2021). Although our results indicate a lower case incidence in children compared to adults, other recent studies based on seroprevalence rates revealed a different picture with comparable seropositivity rates between children and adults (Hyde, 2021). The lower number of cases in children may reflect biases in the exposure and the methodology behind COVID-19 testing (Hyde, 2021). More specifically, the discrepancies between PCR versus serology-identified cases in the pediatric population could be explained by various factors such as a shorter detection window for PCR testing (Lewis et al., 2021) and higher proportion of asymptomatic cases associated with more stringent criteria for children to be tested (Waterfield et al., 2021). This may explain why children were often under-diagnosed and why the role of children in the transmission chain is still under debate.

During the early phase of the pandemic, various public health measures (PHMs) have been implemented to contain the spread of the virus, including curfews and school closures, and as illustrated in Figure 1, the epidemiological curves showed general trends punctuated by some peaks that may be influenced by such public health measures. Further analysis comparing or associating the epidemiological curves and the different non-pharmaceutical measures implemented by each country was not possible due to the low number of reported cases and different measures implemented concurrently. Nevertheless, concerns about their impact on children’s rights to health, social care, and education have been raised from the scientific community. In an extensive review based on major pediatric diseases and referenced modeling projections, Coker et al. (2020) highlighted potential dreadful effects such as increasing severe malnutrition rates, interruption of infectious diseases prevention programs such as vaccination campaigns or long-lasting insecticidal net distribution, and the rise in domestic violence due to the redirection of resources and implementation of public health measures to stop the spread of COVID-19 (Coker et al., 2021). The UNFPA also published a worrying report on the disruption in meeting family planning needs and the increase of gender-based violence and child marriage (UNFPA, 2020).

### Strengths and limitations

To our knowledge, this is the largest analysis on individual-level pediatric COVID-19 cases in Sub-Saharan Africa. Data were presented at three levels: national, regional and in Sub-Saharan Africa as a whole. However, due to constraints on completeness and quality of available data, we could only include data of 15 countries. The cumulative incidence rates differed significantly between countries ranging from 0.2 in Chad to 94.2 in Eswatini. The large differences may partly be explained by differential underreporting of cases and different testing criteria between countries. Nguimkeu et al. (2021), noted that underreporting may be explained by an insufficient diagnostic capacity in certain regions in Sub-Saharan Africa (Nguimkeu and Tadadjeu, 2021). Available data from our focus countries also reflected only few deaths among children, if any. COVID-19 responses in Sub-Saharan Africa have benefited from dynamic, comprehensive, and timely efforts from governments, often in collaboration with various partners.

### Inferences for health policy and governance

The COVID-19 pandemic has highlighted gaps in data management in health systems across the world. Experiences from the pandemic have evidenced the need for effective, efficient, responsible, and participatory governance to ensure children’s health data is used responsibly and not misused (Ienca and Vayena, 2020). Inferences from this research strengthen recent advocacy calls on the urgent need for sex- and age-disaggregated data across countries and regions (Heidari et al., 2020). Recognition of the importance of high-quality data is crucial for impactful analysis on the effect of the COVID-19 pandemic on children in sub-Saharan Africa. While effective and high-quality data collection drives policy, lessons from the pandemic have stressed the need for cross-sectoral trust and collaboration (Zar et al., 2020). National policy and data governance to improve health outcomes among children can benefit from trusted data sharing platforms that enable equitable, inclusive, cross-border data-driven collaborations. This research project strongly recommends key multi-sector stakeholders with varied mandates and interests in sub-Saharan Africa to use the pandemic as an opportunity to collaboratively identify and implement innovative solutions to protect the long-term wellbeing of the region’s children.

## Conclusion

Both the reported cumulative incidence proportions, as well as the number of deaths among the pediatric population studied was smaller than in other age groups such as the adult population. These differences can reflect biases in the exposure and the methodology of the current COVID-19 testing protocols implemented by countries. Out of the under 18 age group, adolescents had the highest cumulative incidence proportion and males were slightly more affected than females. Nevertheless, the underreporting of cases in different regions cannot be ruled out. Even though morbidity and mortality were relatively low in children, negative effects brought by the implementation of public health measures and the redirecting of funds towards the fight against the COVID-19 pandemic have led to an indirect impact on child health. Such changes require a separate and more extensive investigation to provide a more holistic understanding of the effects of the current pandemic on children’s health and lifestyle.

## Data Availability

The data that support the findings of this study are available on request from the corresponding author. Some of the data are publicly available through situation reports produced by Ministries of Health and WHO/AFRO on their respective websites.

## Author Contribution

SRV, LJ, PS, ZH, SBM, AV conceived and designed the study. CBH, CC, OK, and BI made substantial contributions in reviewing the design of the study. JD and BN acquired, managed, cleaned, and analyzed the data. SRV, LJ, PS, and ZH interpreted the data. LJ, PS, and ZH conducted the literature review. SRV, LJ, PS, ZH, AV, and JD drafted the manuscript. CC, CBH, OK, and BI contributed by revising the manuscript critically for important intellectual content. CBH, OK, BI, FM, GSW, NN, AT, ASG, and JWC critically reviewed the manuscript. All authors contributed to final approval of the version to be submitted.

## Acknowledgments

We would like to express our sincere appreciation and gratitude to the entire team at the GRAPH Network and the WHO Regional Office for Africa for guiding and supporting us along the entirety of the study. Additionally, we would like to thank the governments of Botswana, Burkina Faso, Chad, Congo, Eswatini, Guinea, Liberia, Mauritius, Mozambique, Namibia, Niger, Rwanda, Sao Tome e Principe, Sierra Leone, and Uganda for their contribution to the merge line listing dataset.

## Ethical Approval

Data were collected for surveillance purposes under IHR, and no ethical approval was needed.

