## Supplemental Figure 1 for "The toll of COVID-19 on African children: A descriptive analysis on the COVID-19-related morbidity and mortality among the pediatric population in Sub-Saharan Africa"

**Supplementary Material**

Among 47 WHO AFRO countries, only 34 provided data disaggregated data by age and sex to WHO and us. Out of the 34 countries, 13 did not meet the data quality as one country underreported their clinical outcomes, one country underreported their cases, one country had outdated data, and 10 countries had low quality data. Out of the remaining 21 countries, only 15 met the inclusion criteria (Botswana, Burkina Faso, Chad, Congo, Ewsatini, Liberia, Mauritius, Mozambique, Namibia, Niger, Rwanda, Sao Tome Principe, Sierra Leone, and Uganda).

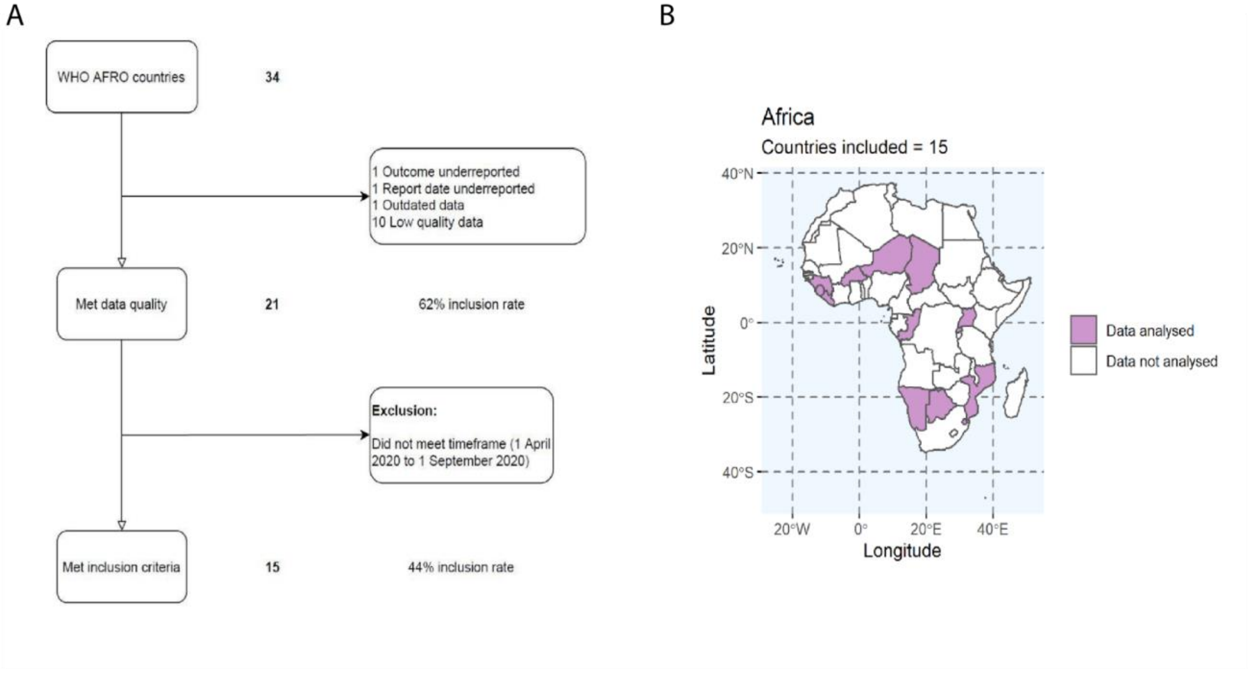


**FIGURE 1.** A) Flowchart for selection of countries to be included in final data analysis; B) Map of Africa highlighting the countries selected for the final data analysis
